# Exploring innovative G-CSF schedules in AML cytarabine-based consolidation through a digital twin study of white blood cell recovery

**DOI:** 10.1101/2025.07.17.25331695

**Authors:** Adrian-Manuel Reimann, Enrico Schalk, Dimitrios Mougiakakos, Thomas Fischer, Sebastian Sager

**Affiliations:** Department of Mathematics, Otto von Guericke University, Magdeburg, Germany; Department of Hematology, Oncology and Cell Therapy, Medical Faculty, Otto von Guericke University, Magdeburg, Germany; Institute of Molecular and Clinical Immunology, Medical Faculty, Otto von Guericke University Magdeburg, Magdeburg, Germany; Healthcampus Immunology, Inflammation and Infectiology (GC-I3), Otto von Guericke University Magdeburg, Magdeburg, Germany; Center for Health and Medical Prevention – CHaMP, Otto von Guericke University Magdeburg, Magdeburg, Germany; Max Planck Institute for Dynamics of Complex Technical Systems, Magdeburg, Germany

**Author notes:** Corresponding Author: Adrian-Manuel Reimann.

**Keywords:** Acute Myeloid Leukemia, Digital Twins, Mathematical Modeling, Chemotherapy, Myelosuppression

## Abstract

Patients with acute myeloid leukemia (AML) are at high risk for life-threatening infectious complications due to chemotherapy-induced leukopenia. In contrast to other malignancies, prophylactic administration of granulocyte colony-stimulating factors (G-CSF) is not routinely done in AML. However, recently, clinical trials showed that administration of G-CSF post consolidation chemotherapy leads to faster white blood cell (WBC) recovery and to fewer infections. We investigated novel G-CSF schedules using a mathematical model trained with retrospective clinical data (digital twins) consisting of high-frequency measurements of 65 patients. We evaluated 323 different treatments varying in start and duration of G-CSF administration by examining the impact on the occurrence and duration of leukopenia. For each treatment, we used the same 65 digital twins, plus an additional 100 artificial patients for uncertainty quantification, receiving 3 consolidation cycles each. We lumped the 323 treatments into 4 clusters: NO-G-CSF, PRE-G-CSF, SIM-G-CSF, and POST-G-CSF for schedules without G-CSF, that terminated G-CSF before consolidation chemotherapy started, that overlapped on at least one day, and that started post chemotherapy, respectively. The numerical simulations resulted in substantial differences in the occurrence of leukopenia: 88% of all consolidation cycles with leukopenia for NO-G-CSF, 98% for SIM-G-CSF, 45% for POST-G-CSF, and 28% for PRE-G-CSF. Administration of G-CSF prior to chemotherapy may thus significantly enhance the efficacy of AML consolidation therapy and therefore warrants clinical investigation.

## INTRODUCTION

The primary goal of therapy in patients with acute myeloid leukemia (AML) is remission induction. In general, after achieving remission, patients with a relatively low recurrence risk of <35-40% [1] receive consolidation with intermediate-dose (ID) or high-dose (HD) Ara-C [2] which as a drug disrupts DNA synthesis, particularly in rapidly dividing cells, whereas high-risk patients undergo hematopoietic stem cell transplantation. Depending on the molecular/cytogenetic profile, the addition of other agents in induction and consolidation, such as midostaurin for FLT3-mutated patients [3,4], was shown to improve prognosis. Patients with AML are at increased risk for developing life-threatening infectious complications, such as pneumonia or bloodstream infections, both as a result of AML-induced neutropenia and intensive treatment with cytotoxic (chemo-) therapy [5]. Patients with AML have a very high risk (>80%) of infections, which are the primary cause of death during therapy in >70% of cases and are one of the most common reasons for transfer to the intensive care unit [6,7].

Neutropenia, defined as an absolute neutrophil count of <500/µL, or in general, leukopenia characterized by a white blood cell [WBC] count <1000/µL as a surrogate [8], is the main risk factor for infectious complications in patients with AML [9]. The incidence and severity of infections depend on the depth and duration of neutropenia [5,10]. In patients with AML, microbiologically documented infections tend to occur at the time point of the neutrophil nadir, while clinically documented infections generally occur some days earlier [11]. Another important risk factor of infection is the degree and timing of gut mucosal damage. Neutropenia aggravates this complication and may result in the clinical presentation of neutropenic enterocolitis [12].

For some non-myeloid malignancies with high-dose chemotherapy treatments, administration of granulocyte colony-stimulating factors (G-CSF) is recommended, however, the most effective timing is still under discussion [13]. In comparison, prophylactic administration of G-CSF is not generally recommended in AML [14], although it is capable of reducing both the duration and the severity of neutropenia [15]. It remains controversial due to a theoretically increased risk of relapse and indeterminate impact on neutropenic complications [16]. Results from clinical trials showed that in AML consolidation therapy, administration of Ara-C on days 1, 2, and 3 (Ara-C-123) was superior in comparison to Ara-C on days 1, 3, and 5 (Ara-C-135) because of faster hematologic recovery and a lower incidence of infections. Interestingly, pegfilgrastim, a depot G-CSF formulation, when administered a few days after the consolidating chemotherapy, further reduced the infection rate [17,18]. However, improved survival was not seen [17,18].

In addition, previously, G-CSF has been applied concurrently with AML induction chemotherapy as a priming approach to drive leukemic stem cells into S-phase of the cell cycle. This approach aimed to increase the sensitivity to S-phase specific chemotherapy such as cytarabine. However, the results of several randomized trials did not justify further application of this strategy [19]. For example, neutrophils regenerated with a delay (P = .007) in patients assigned to a concurrent G-CSF treatment regimen [20]. Also in our simulations, the concurrent, overlapping administration of cytarabine and G-CSF during consolidation therapy did result in delayed WBC recovery times. We considered novel G-CSF schedules that, like priming, are applied before consolidation chemotherapy, but do not overlap with induction chemotherapy. We hypothesized that in AML consolidation therapy, such innovative G-CSF schedules have the potential to substantially reduce the nadir and the duration of leukopenia and may significantly improve the outcome of AML treatment. Therefore, we investigated novel G-CSF schedule variants and examined the induced changes in the occurrence and duration of leukopenia upon AML consolidation therapy. Motivated by the large number of existing G-CSF schedules, we used digital twins, specified in the upcoming section and the supplementary material, to fully explore the wide spectrum of different combinations regarding both the starting point of administration and the duration of G-CSF application.

## MATERIALS AND METHODS

Referring to the different possible concepts for digital twins in oncology [21], we define a digital twin as a mechanistic mathematical model with model parameters that allow personalization with clinical data. In this study, we used 65 digital twins that are specified via six individual model parameters each in the supplementary material. The training data were obtained retrospectively from the phase II AMLSG 12-09 randomized-controlled trial (RCT; L=44), collected between 2010 and 2012 [22] and from clinical chart records obtained between 2008 and 2015 from the Magdeburg University Hospital, Germany (L=21). We had no access to information that could identify individual participants from the phase II AMLSG 12-09 trial. Data from the Magdeburg University Hospital was pseudonymized. The group was heterogeneous considering Ara-C administration and timing, whether or not and how lenograstim as a G-CSF was administered, age (median 58.5 years), and number of observed longitudinal WBC counts, with a total of 1869 observations. Variance of G-CSF schedules was not a focus of AMLSG 12-09, and none of the patients received a non-standard PRE-G-CSF or SIM-G-CSF treatment.

Digital twins are able to predict, within a margin of uncertainty, outcomes of different treatments. Prediction of outcomes for treatments that were not covered in the training data (like different G-CSF schedules) is challenging. Therefore, we used a mechanistic model including pharmacodynamics and pharmacokinetics specified in our supplementary material. The relatively high number of approximately 30 observed time points per patient distinguishes this data set from other data sources in AML and was the reason why we used it for the training of digital twins. In a previous study [23], it was shown that the distributions of WBC recovery times of this cohort with 65 patients did not differ significantly from those of larger clinical cohorts, also for treatments that were only partly included in the training set. The study in [24] showed that digital twins using the aforementioned model and data were even capable of reproducing individual treatment outcomes. The underlying system of differential equations, more background on the retrospective, longitudinal data containing WBC count measurements, treatment information, measurements of leukemic blasts in the bone marrow, and uncertainty quantification via propagation of random variations of model parameters to performance indicators of interest are specified in our supplement.

In addition to the mathematical model with personalized model parameters specifying a digital twin, three more clarifications are necessary. In “Treatment choices” we discuss the degrees of freedom associated with the clinical treatment, i.e., the administration of Ara-C and G-CSF. In “Considered scenario” we specify the length of the simulation horizon and our choice of the initial values of all differential states. In “Simulating and predicting treatment outcomes” we explain how we evaluated WBC recovery based on the numerical solution of differential equations (simulation). In summary, our approach allows us to specify G-CSF schedules (at what times, how much is administered) and obtain corresponding WBC recovery times and blast ratios for all simulated CCs of all digital twins.

### Treatment and schedule choices

In the interest of a clear focus on different G-CSF schedules, chemotherapy treatment was specified to the administration of high dose 3 g/m^2^ (n=30) or intermediate dose 1-1.5 g/m^2^ (n=35) Ara-C on days 1, 2, and 3 of each consolidation cycle (CC), with the dosage chosen according to the underlying clinical data to reduce extrapolation errors of the mathematical model. Throughout, as it did not affect the qualitative result, a BSA of 2 m^2^ was assumed. Ara-C was administered twice a day at 8 am and 8 pm consistently across all simulations. The length of all CCs was fixed to 35 days. The length of CCs has an important impact on WBC recovery time [23]. Yet, for each choice we considered other than 35 days, the results concerning relative improvements due to G-CSF administration were qualitatively similar, as was also the case for Ara-C135 (data for both settings not shown). We fixed the dosage of G-CSF to one infusion of 263 µg per day at 12 pm. We focused on different timings and lengths of G-CSF administration. We defined the starting point “s” of G-CSF treatment relative to the first CC day on which the first dosage of Ara-C is administered: G-CSF starting day 0 indicates a simultaneous start of G-CSF and Ara-C treatment on the first day of each CC. We varied the starting day of G-CSF treatment from -23 to 21 in steps of 2 days and the duration “l” of treatment from 1 to 14 in steps of 1 day. Adding no G-CSF treatment (i.e., l=0), this resulted in n=23×14+1=323 treatments.

To be able to assess the question if administration of G-CSF before, during, or after the administration of chemotherapy is beneficial in comparison to not giving G-CSF, we clustered the 323 treatments in 4 classes. Using s as start day and l as duration length, we defined POST-G-CSF as the union of all treatments with s>=3 (140 schedules), PRE-G-CSF as the union of all treatments with l+s<1 (119 schedules), and SIM-G-GCSF as the union of all remaining treatments (63 schedules). NO-G-CSF contains only 1 schedule, the one with l=0.

### Considered scenario

We considered a time horizon of 154 days, consisting of a first interval of 49 days, followed by three CC of 35 days each. The first period of 49 days was used to simulate the effect of a previous chemotherapy treatment (such as induction therapy) on the differential states. This scenario was meant to reflect the clinical setting in which consolidation therapy follows upon induction therapy, and the hematologic system can be expected to be in a phase of recovery. We also considered a scenario without this previous chemotherapy treatment, where all initial values started from a steady state (data not shown, results were qualitatively similar). The concept is visualized in Figure 1. In Figure 2C, a prototypical WBC trajectory is shown over the time period from day 1 to 154 for G-CSF treatments with three different administration timings, which are also plotted.

**Caption Fig. 1.**
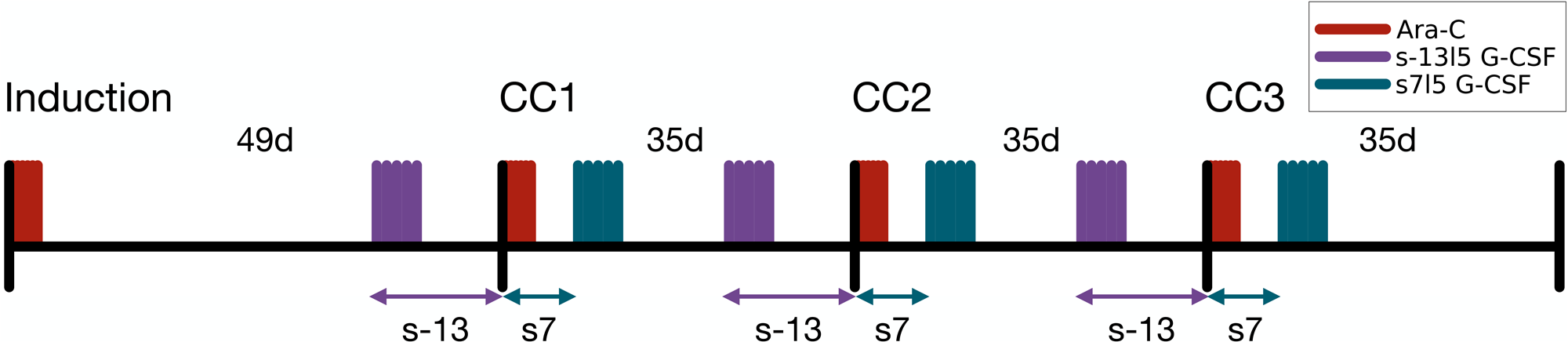
Illustration of the simulation specifics. The first interval of 49 days allows the consideration of G-CSF treatments starting before Ara-C treatment, i.e., before a consolidation cycle (CC) begins. This is necessary for some of the treatments with the start of G-CSF s<0. Considering the time interval of 154 days, Ara-C is always administered on days 1, 2, 3 (induction), 50, 51, 52 (CC1), 85, 86, 87 (CC2), 120, 121, and 122 (CC3). The G-CSF administration depends on the specifications of s and l. For example, for s=-13 and l=5, it is administered on days 37-41 (CC1), 72-76 (CC2), and 107-111 (CC3).

**Caption Fig. 2.**
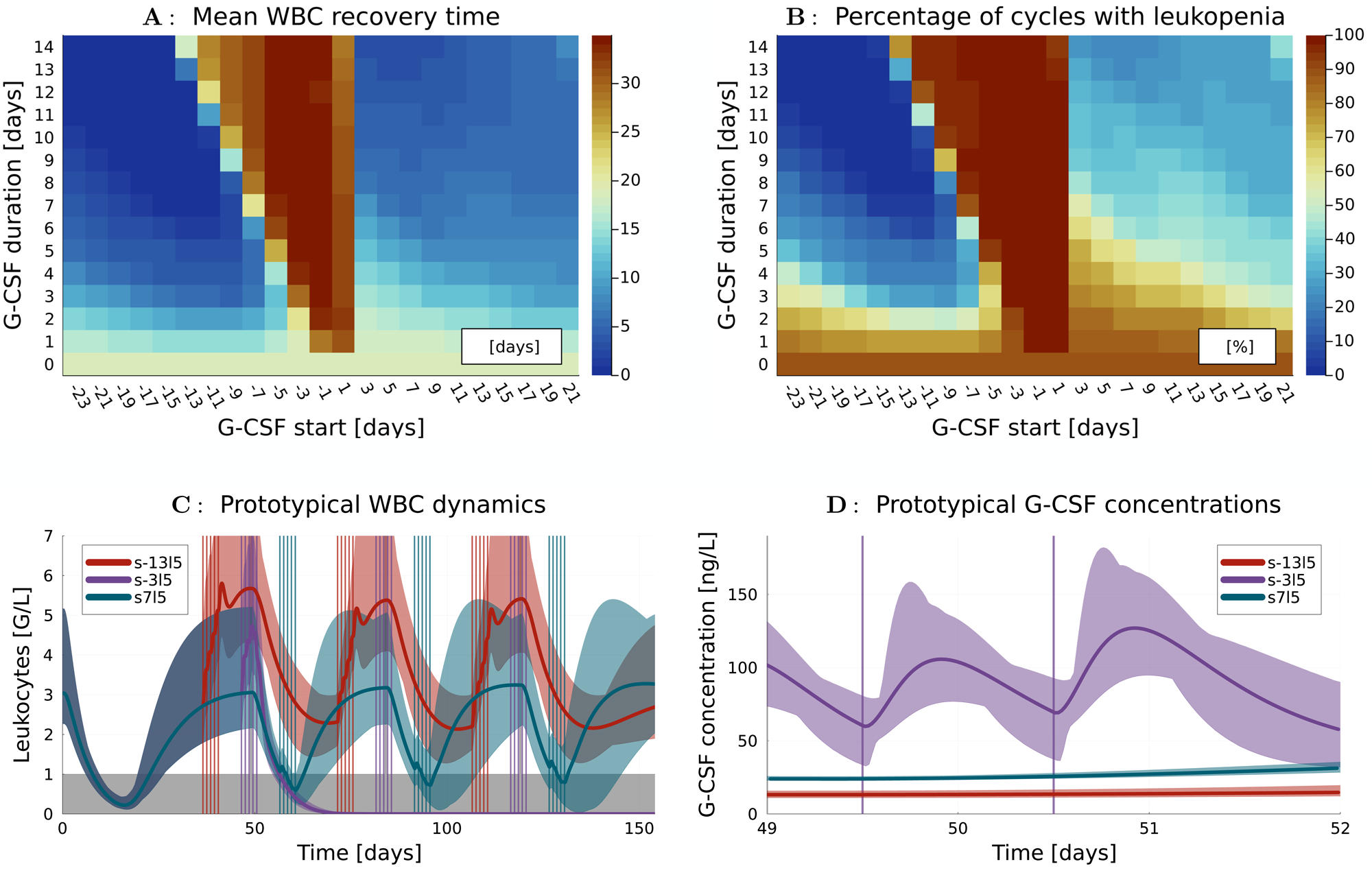
Simulated WBC recovery after chemotherapy and G-CSF treatment. **A** The heatmap illustrates the WBC recovery time in days for different G-CSF schedules. They differ in the start s of the treatment (x-axis) relative to the beginning of Ara-C administration and in the number of consecutive days G-CSF is administered (G-CSF duration l on the y-axis). E.g., a G-CSF start s=-3 denotes G-CSF administration starting three days before Ara-C administration, and a duration of l=5 denotes that G-CSF is administered for 5 consecutive days. The color of each cell encodes mean values taken over 195 simulated CC of 65 digital twins for a particular G-CSF treatment. The mean is lowest (blue) for treatments on the left hand side of the plot, corresponding to early administration of G-CSF before chemotherapy. The mean is slightly higher (light blue) on the right hand side of the plot, corresponding to G-CSF treatments after termination of chemotherapy. The G-CSF treatments overlapping with chemotherapy show the worst performance of more than 30 days (red). In addition, a general trend that longer durations lead to faster hematologic recovery can be observed. **B** A similar heat plot shows a color encoding of the percentages of the 195 CCs for each G-CSF treatment, in which a leukopenia occurred. The heat plot has a similar geometry to the one shown in Figure 2A, with a more emphasized difference between the percentages close to 0% on the left hand side and the percentages ranging between 30% and 90% on the right hand side. **C** For a prototypical digital twin and three exemplary G-CSF treatments of the same duration l=5, the G-CSF timing (vertical bars) and the corresponding time trajectories of WBC counts, together with an uncertainty range as specified in the supplementary material, are plotted. The red trajectory corresponding to the early G-CSF treatment s-13l5 oscillates between values well above the critical leukopenia region in grey. The violet trajectory corresponding to a G-CSF treatment overlapping with chemotherapy s-3l5 temporarily increases the WBC count, but shows a drastic reduction of WBC, from which it does not recover in the simulated time horizon. The blue trajectory corresponding to G-CSF after Ara-C, s7l5, shows temporary increases of WBC counts, but also time periods of leukopenia. **D** Corresponding G-CSF concentrations are plotted for days 49-52 when chemotherapy is administered. The increased G-CSF values for s-3l5 result in a stronger impact of Ara-C on the decrease of WBC observed in Figure C.

### Simulating and predicting treatment outcomes

We calculated treatment outcomes for all digital twins by simulation using the *RosenbrockW6S4OS* method from *DifferentialEquations.jl*. We used the WBC recovery time for every CC as a key performance indicator for leukopenia severity. WBC recovery time is a clinically established parameter.

In summary, we obtained results for 323 different G-CSF treatments and 65 digital twins, and 3 consecutive CCs, each. For all we extracted the WBC recovery times. The WBC recovery time was set to 0 if the WBC was above 1000 /µL at all times, i.e., if no leukopenia occurred.

## RESULTS

To illustrate the distribution of WBC recovery times, Figure 2A shows the mean (concerning the 195 CCs of 65 digital twins) WBC recovery times resulting in one value for each of the 323 different G-CSF treatments. Each cell in the heat plot corresponds to a particular G-CSF treatment, specified via starting day s on the horizontal axis and duration length l on the vertical axis, and corresponds to the mean WBC recovery time indicated by the heatmap.

One can visually discern three distinct classes among the 322 G-CSF treatments, aligning with the pre-defined sets PRE-G-CSF, SIM-G-CSF, and POST-G-CSF. SIM-G-CSF, the triangular-shaped area with many red cells, shows the poorest performance concerning WBC recovery time, even worse than NO-G-CSF (the cell with l=0). The POST-G-CSF treatments with starting day s>=3 correspond to the light bluish area on the right-hand side of the red triangle. A good choice here is s=l=7, resulting in a mean WBC recovery time of 6 days. The PRE-G-CSF treatments correspond to the dark bluish area on the left-hand side of the red triangle. An exemplary good choice for PRE-G-CSF is s=-13, l=7, resulting in a mean WBC recovery time of 0 days.

Figure 2B shows in a similar way the percentages of leukopenic CCs, i.e., for which the WBC was at least at one time point below 1000 /µL. The visual separation in the heat plot is identical to Figure 2A. Again, SIM-G-CSF treatments form a triangle, affecting almost all digital twins with up to 100% leukopenia occurrences. The situation is drastically better for POST-G-CSF treatments, which reduce the incidence of leukopenia to percentages between 30% and 90%, approximately. A clear trend indicates that the longer G-CSF is administered, the lower the occurrence of leukopenias, with decreasing percentages observed towards the upper end of the subplot. This lowering risk of leukopenias is mostly due to the strong increase of WBC that carries over to the next simulated CC, resulting in the Chemotherapy not lowering the WBC below 1000 /µL after the first CC. The PRE-G-CSF treatments have the potential to reduce the percentage of leukopenias to 0%, e.g., for s=-13 and l=7 and longer durations since the overlapping effect observed in the top right area for POST-G-CSF is already present for the first CC when considering PRE-G-CSF.

To quantify these visual impressions, we also looked at mean values for all treatments in the four classes. The numerical simulations resulted in different outcomes concerning mean WBC recovery times in days and percentage of CCs with leukopenia: 19 days and 88% for NO-G-CSF, 32 days and 98% for SIM-G-CSF, 8 days and 45% for POST-G-CSF, and 6 days and 28% for PRE-G-CSF.

To get a better understanding of the WBC recovery in response to the different treatment schedules, Figure 2C shows for one prototypical digital twin and an uncertain (see supplementary material for details) evolution of WBC counts over time for three different G-CSF application schedules. The length of all treatments is identical with l=5 and the starting days s=-13, s=-3, s=7 are shifted by 10 days, such that we have one example from the PRE-G-CSF, SIM-G-CSF, and POST-G-CSF group, respectively. The early administration of G-CSF at s=-13 leads to oscillations in the range between 3000 /µL and 6000 /µL. For starting date s=-3 with an overlap between G-CSF and Ara-C administration, the WBC count drops dramatically. For the schedule to administer G-CSF after Ara-C, here at s=7, we observe a temporary increase of WBC counts, which does not result in completely avoiding leukopenia, though.

Figure 2D zooms into the critical time horizon between days 49 and 52 when Ara-C is administered and visualizes the concentration of G-CSF in the body. One observes that, due to the administration of G-CSF, the concentration for s=-3 is significantly higher than for the s=-13 and s=7 treatments. Thus, the increased G-CSF exposure at a time point when Ara-C effects peak leads to increased hematopoietic toxicity and the delayed but steep decline of WBC shown in Figure 2C.

## DISCUSSION

### Main results

Our main hypothesis was that novel schedules of G-CSF application employing early administration prior to consolidation chemotherapy might result in fewer and less severe leukopenias in AML. Leukopenia in AML is generally viewed as a measure of neutropenia [8]. As apparent from Figures 2A and 2B and from the mean values of 28% CCs with leukopenias for PRE-G-CSF in comparison to 45% for POST-G-CSF, 98% for SIM-G-CSF, and 88% for no G-CSF at all, this hypothesis seems plausible.

The advantage of early G-CSF administration is not a simple consequence of more time for WBC to grow, but of more complex dynamics. This can be seen by looking at the oscillatory nature of the system, which would result in WBC going back to a steady state value in the long run (compare Figure 2C), and by the non-monotonicity observable for the SIM-G-CSF schedules.

A secondary result is that for both PRE-G-CSF and POST-G-CSF treatments, longer durations provide better results with respect to WBC recovery. Disadvantages of prolonged G-CSF administration, such as increased risk of adverse effects or economic costs, are not considered here.

### The study’s rationale

Although classical consolidation therapy has been based on Ara-C for decades, there are open questions concerning the optimal number and duration of cycles as well as dosage and timing of Ara-C administration [17,18,25]. If and how G-CSF should be prophylactically administered remains an open question and is not generally recommended in patients with AML [14]. An examination of US American guidelines [26] references the FDA label stating that pegfilgrastim should be avoided 14 days before or within 24 hours of administration of myelosuppressive chemotherapy [27] and attributes this to the theoretical potential to increase the toxicity of chemotherapy to myeloid progenitor cells following growth factor stimulation [28]. Studies reported a strong increase in the incidence of severe neutropenia in In general, caution is warranted when comparing different G-CSF formulations, such as lenograstim and pegfilgrastim, as potential differences in pharmacokinetics, dosing schedules, and clinical effects cannot be excluded. However, our findings suggest that certain similarities in their behavior may be inferred. For example, we observed an increase in the number and severity of leukopenias, also in our simulations. These outcomes are well matched by the red triangles in Figures 2A and 2B, corresponding to SIM-G-CSF, and are plausible by looking at the physiological properties encoded in the mathematical model. Large G-CSF concentrations stimulate cell proliferation, which leads to a strong increase in chemotherapy toxicity when administered at the same time. The recommended 24h margin aligns well with our results for s=3. However, our results (left-hand side of the red triangle) show that a safety distance of 2 days is enough when comparing the end of G-CSF administration and the beginning of chemotherapy. This gives ample room for treatment improvements when compared to the recommended 14 days [26].

Regarding the potential for additional prophylactic G-CSF administration applied after chemotherapy, there are contradictory results in the literature. Only minor improvements in the depth and duration of nadir and thus in infection and mortality rates have been reported [31,32]. In contrast, recent clinical studies [17,18] and a study based on digital twins [23] reported a significant shortening of WBC recovery times of approximately 2 days due to G-CSF administration after chemotherapy.

For other malignancies, the timing of G-CSF is also under current debate. E.g., for patients with lung cancer receiving chemotherapy weekly, the question of same-day versus next-day administration of G-CSF has been discussed [33,34]. Note that this discussion corresponds to a discussion of the exact boundary between SIM-G-CSF and POST-G-CSF in our setting, i.e., to the vertical lines around s=3 in Figures 2A and 2B. The result that both seem to have a similar, beneficial impact on the reduction of febrile neutropenia [33,34] is consistent with our simulation results. Note that logistical aspects of different G-CSF administrations have also been discussed in this context [35]. However, direct comparison to the AML setting should be made with caution, as treatment response, underlying mechanisms, and treatment guidelines generally differ. If increased duration or dosage of G-CSF is considered, corresponding to better performing upper rows in Figures 2A and 2B, possible side effects of overdosage [36] and economic and logistic costs also need to be considered.

### Choice of methods

There are different purposes and ways of using digital twins [21]. Here, we decided to use a mechanistic mathematical model that was trained and cross-validated with clinical data and used (partially) in a series of previous publications [23,24,37–39]. It focuses on aspects that are relevant for the open questions mentioned above, i.e., myelosuppression, pharmacokinetics, and -dynamics of Ara-C and G-CSF (lenograstim). Advantages in comparison to deep learning-based approaches are interpretability, better extrapolation properties, and a reduced amount of necessary training data. The simulated outcomes are, of course, uncertain and have to be verified by clinical studies. The fact that none of the 65 patients at the origin of the training data received a non-standard PRE-G-CSF or SIM-G-CSF treatment may impact the reliability of the mathematical model for these treatments. High dose chemotherapy is a large perturbation of the bone marrow microenvironment, the hematopoietic cells, and the cytokine feedbacks. Therefore, it is not clear if the implied assumption that hematopoietic cells before, during, and after chemotherapy respond identically to external G-CSF administration is valid. Therefore, all results need to be interpreted very carefully. Yet, our goal was not to predict individual responses of individual patients as well as possible, but rather to obtain a representative cohort of patients, in which such uncertainties might also balance out. The smoothness of the results with respect to G-CSF starting date and duration in Figure 2, also for the supporting uncertainty quantification results in the supplement, makes it, in our opinion, a clear indication that it is worthwhile to follow these indications with a validating clinical trial.

### Clinical outlook

We propose to explore the investigated early administration of G-CSF in a clinical trial or a mouse clinical trial. We anticipate that our results based on digital twins will translate qualitatively into clinical reality. Initiating G-CSF administration early in the treatment course may enhance the therapeutic effectiveness of AML consolidation therapy.

However, improvements of classical consolidation therapy have to be seen in the context of the trend to an increase in allogeneic hematopoietic stem cell transplantation after induction therapy and to use targeted therapies involving, e.g., FLT3-or IDH-inhibitors. A development of digital twins able to represent these treatments might help to design even better leukemia therapies. For the future, we believe that investigation of novel G-CSF schedules in digital twins may offer major improvements in WBC recovery and thus in hematologic toxicity in a wide variety of chemotherapy schedules of other hematologic and solid malignancies.

## Supporting information

Supplemental Materisl

## Acknowledgements

We sincerely thank Prof. Schlenk et al. and the University Clinic Ulm for kindly providing the data obtained in the AMLSG 12-09 trial that has been crucial for this study.

## Author Contributions

SS designed the study; AR implemented numerical algorithms and visualized results; ES and TF contributed to data and result interpretation. All authors contributed to the writing of the manuscript. They read and agreed on the final version of the manuscript.

## Data Availability Statement

A complete specification of all 65 digital twins is provided in the online supplement. All results should be reproducible up to numerical accuracy based on the differential equations, the specification of initial values, and these model parameters. Further enquiries can be directed to the corresponding author.

## Statement of Ethics

This study purely builds on computational results using data from two sources that have already been published. Written informed consent was obtained from the patients treated within the AMLSG 12-09 trial; the corresponding protocol was approved by the central ethics committee at the time [22]. The second study used was approved by the ethics committee of the Magdeburg University Hospital (approval no. 124/15). Given the nature of a retrospective chart review study and using routine clinical data, written informed consent was not required within the study (ethics committee Magdeburg University Hospital, approval no. 124/15).

