## Supplemental Materisl for "Exploring innovative G-CSF schedules in AML cytarabine-based consolidation through a digital twin study of white blood cell recovery"

**Supplemental Material**

This supplemental material complements the research article *Exploring innovative G-CSF schedules in AML cytarabine-based consolidation through a digital twin study of white blood cell recovery*. It makes relevant data available. To ensure that the manuscript together with this supplementary document is self-contained, we also include material that was already published before in [Reimann et al., 2023], [Jost et al., 2020], or [Jost et al., 2019]. Parts of this supplement are identical copies of content of the supplement to [Reimann et al., 2023].

We provide details concerning data in Sections 1 and 2. A detailed description of the mathematical model follows in Section 3. Parameters for this model and details how they were derived are presented in Section 4. We discuss key performance indicators for leukopenia in detail in Section 5. This section also includes an uncertainty quantification of the main result via propagation of uncertain model parameters via the dynamic system to the key performance indicators. Finally, also the impact of the treatment schedules on predicted blast growth is investigated in Section 6.

### 1 Training Data

We trained 65 digital twins using longitudinal retrospective data. The data used was obtained from the phase II AMLSG 12-09 randomized controlled trial (RCT; $n = 44$) and from clinical chart records from the Magdeburg University Hospital, Germany ($n = 21$). This study was approved by the ethics committee of the Magdeburg University Hospital, approval no. 124/15. The group was heterogeneous considering Ara-C administration, lenograstim administration, and age.

Table 1: Numbers and age of patient subgroups in the training data

| Data source | Magdeburg | | | Ulm | | Sum |
|---|---|---|---|---|---|---|
| Ara-C | HDAC-123 | HDAC-135 | IDAC-135 | HDAC-123 | IDAC-123 | all |
| #patients | 1 | 11 | 10 | 18 | 25 | 65 |
| with blast measurements | 0 | 0 | 0 | 15 | 20 | 35 |
| with lenograstim treatment | 0 | 0 | 0 | 13 | 22 | 35 |
| median age [y] | 48 | 62 | 63 | 55.5 | 72 | – |
| mean age [y] | 48 | 62.8 | 54.9 | 51.6 | 70.5 | – |

Details are shown in Table 1. The data set is identical to the one of a previous study [Jost et al., 2020], but for two patients who have been removed for reasons of data inconsistency, as in [Reimann et al., 2023]. The data contains longitudinal patient-specific data, most importantly 1869 white blood cell (WBC) concentration measurements and 63 relative blast measurements from the bone marrow.

### 2 Discussion of Appropriateness of the Training Data for the Virtual Study

Note that the focus of the clinical studies mentioned above was not on an investigation of early administration of G-CSF. The concurrent administration of cytarabine and G-CSF allowed us the calibration of our mathematical model, though. The time series were used to estimate model parameters, which allows an extrapolation of predictions towards novel treatment schedules as the one investigated in our manuscript. A clinical verification of the predicted results was thus not a priori available nor at the basis of the mathematical model. While it is possible, or even likely, that the mathematical model does not capture all aspects of PRE-G-CSF treatments and there might be structural, epistemic uncertainties involved, those can only be overcome by actually running a clinical trial. To overcome this chicken-egg problem, we thus suggest a clinical trial as a next step on the way to a change in clinical treatment protocols, and to provide in a synergistic way new data with valuable information for model identification.

### 3 Mathematical Model

There are many levels on which the dynamics of AML treatment can be modeled, see [Clairambault, 2009, Stiehl and Marciniak-Czochra, 2012, Brady and Enderling, 2019, Chulián et al., 2022] for further references. In [Jost et al., 2020], a differential equation model was presented that combines four submodels for hematopoiesis [Friberg et al., 2002, Jost et al., 2019], pharmacokinetics and -dynamics of cytarabine [Jost et al., 2019], leukemic cells [Stiehl et al., 2018], and G-CSF [Jost et al., 2020]. This comprehensive model allows to study in silico the impact of different treatment protocols on WBC (and hence neutropenia depth and length) and on the leukemic blasts.

Figure 1: A visualization of the complete differential equation model

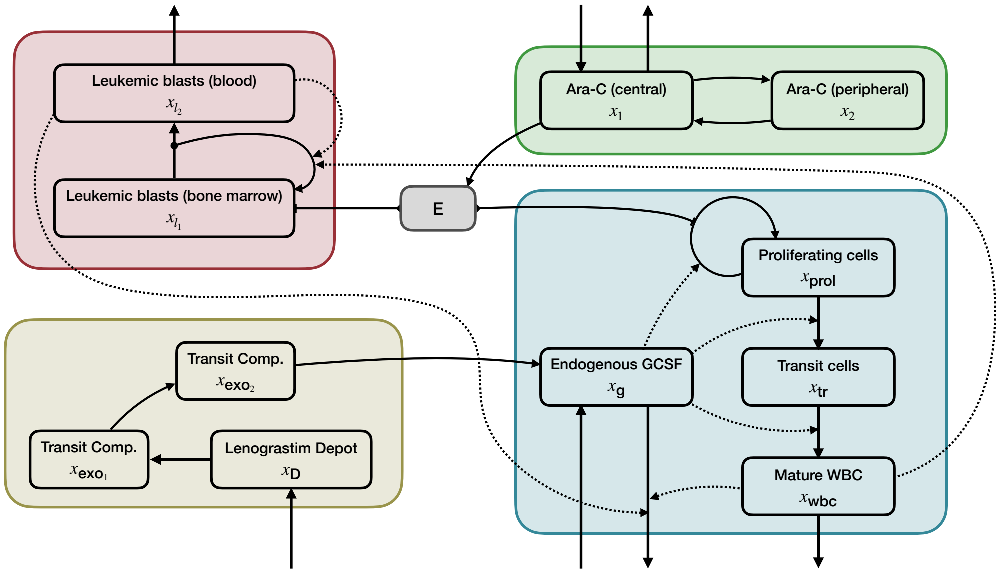


The differential states and functional relations between them are visualized in Figure 1. Shown are differential states $x$ and the pharmacodynamics $E$. Four submodels have been coupled and are indicated in different colors. The myelosuppression compartment model [Friberg et al., 2002] plus endogeuous G-CSF is shown in blue, the leukemic blast compartment model [Stiehl et al., 2018] in red, the pharmacokinetics of cytarabine [Jost et al., 2019] in green and a pharmacokinetics model for lenograstim in yellow. Functional relationships are indicated by arrows connecting the compartments.

The full system of differential equations is

$$\dot{x}_1 = k_{21}x_2 - (k_{10} + k_{12})x_1 + \frac{u_c \text{BSA}}{\text{dur}_c} \tag{1}$$

$$\dot{x}_2 = k_{12}x_1 - k_{21}x_2 \tag{2}$$

$$\dot{x}_{prol} = -\left(\frac{x_g}{B_g}\right)^{\beta} k_{tr}x_{prol} + \left(\frac{x_g}{B_g}\right)^{\gamma S_{x1}} k_{tr}(1-E)x_{prol} \tag{3}$$

$$\dot{x}_{tr} = \left(\frac{x_g}{B_g}\right)^{\beta} k_{tr}(x_{prol} - x_{tr}) \tag{4}$$

$$\dot{x}_{wbc} = \left(\frac{x_g}{B_g}\right)^{\beta} k_{tr}x_{tr} - k_{wbc}x_{wbc} \tag{5}$$

$$\dot{x}_g = k_{in} - k_{out}x_g \tag{6}$$

$$\dot{x}_D = -k_{a1}x_D + \frac{u_l 1000}{V_g \text{dur}_l} \tag{7}$$

$$\dot{x}_{exo_1} = k_{a1}x_D - k_{a2}x_{exo_1} \tag{8}$$

$$\dot{x}_{exo_2} = k_{a2}x_{exo_1} - k_{a2}x_{exo_2} \tag{9}$$

$$\dot{x}_{l1} = (2a_1k_{lc} - 1)p_1x_{l1} - p_1Ex_{l1} \tag{10}$$

$$\dot{x}_{l2} = 2(1 - a_1k_{lc})p_1x_{l1} - d_2x_{l2}, \tag{11}$$

with parameter-dependent initial values specified in Table 2

Table 2: Initial values for all differential states

| **States** | **Initial values** |
|---|---|
| $x_1, x_2, x_{\text{exo1}}, x_{\text{exo2}}, x_D$ | 0 |
| $x_{\text{prol}}, x_{\text{tr}}$ | $(B\ k_{wbc})/k_{tr}$ |
| $x_{wbc}$ | $B$ |
| $x_g$ | $B_g$ |
| $x_{l1}$ | $x^0_{blasts}$ |
| $x_{l2}$ | $B/99$ |

and algebraic variables

$$E = \text{slope}\ \log\left(\frac{x_1}{V_c MM_{AraC}} + 1\right) \tag{12}$$

$$S_{x_1} = 1 + \log\left(\frac{x_1}{V_c MM_{AraC}} + 1\right) \tag{13}$$

$$k_{in} = (k_{e,g} + k_{ANC}B)B_g + k_{a2}x_{exo_2} \tag{14}$$

$$k_{out} = k_{e,g} + k_{ANC}(x_{wbc} + x_{l2}) \tag{15}$$

$$k_{lc} = \frac{1}{1 + c_1x_{wbc} + c_2x_{l2}}. \tag{16}$$

describing the pharmacodynamics of cytarabine $E$, a term $S_{x_1}$ to modeling a secondary effect of cytarabine introduced and discussed in [Jost et al., 2019], terms $k_{in}$ and $k_{out}$ modeling the interactions in the G-CSF compartment, and a term $k_{lc}$ modeling the influence of WBC counts on the proliferation of leukemic blasts [Stiehl et al., 2018].

The mathematical model itself has already been used in previous studies [Reimann et al., 2023, Jost et al., 2019, Jost et al., 2020]. We refer to them for a discussion of modeling assumptions and alternatives and a comprehensive literature survey.

### 4 Parameter Estimation

For most of the differential states in the equations (1-11) no measurements are available, with the exception of WBC counts ($x_{wbc}$) and very few relative leukemic blast measurements. As a consequence, most of the model parameters had to be fixed to values from the literature. Table 3 gives an overview.

Table 3: List of all non–personalized model parameters

| Constant | Unit | Value | Description |
|---|---|---|---|
| **PK model of Ara-C** | | | |
| $k_{10}$ | $1/day$ | 98.2920 | Elimination rate of Ara-C |
| $k_{12}$ | $1/day$ | 2.6616 | Distribution rate of Ara-C |
| $k_{21}$ | $1/day$ | 12.8784 | Distribution rate of Ara-C |
| Volume $V_c$ | $L$ | 26.5554 | Volume of central compartment |
| $MM_{AraC}$ | $g/mol$ | 243.2170 | Molecular mass of Ara-C |
| $dur_c$ | $day$ | 0.1250 | Infusion time |
| **PD model of WBCs and leukemic blasts** | | | |
| $k_{wbc}$ | $1/day$ | 2.3765 | Death rate of circulating WBCs |
| $\beta$ | - | 0.2340 | Feedback of G-CSF on transit time |
| $B_g$ | $ng/L$ | 24.4000 | Endogenous G-CSF steady state |
| $k_{e,g}$ | $1/day$ | 0.5920×24 | Non-specific elimination rate constant |
| $k_{ANC}$ | $1/day$ | 5.6400×24 | Neutrophil-dependent elimination rate |
| $a_1$ | - | 0.8750 | Probability of self-renewal |
| $p_1$ | $1/day$ | 0.1000 | Leukemic cell proliferation rate |
| $d_2$ | $1/day$ | 2.3000 | Leukemic cell death rate |
| $c_1$, $c_2$ | $L/10^9$ | 0.0100 | G-CSF quasi steady-state feedback scaling factor |
| **PK model of lenograstim** | | | |
| $k_{a2}$ | - | $\frac{10}{3}k_{a1}$ | 2. absorption rate of lenograstim |
| $V_g$ | $L$ | 14.5 | Volume of distribution |
| $dur_l$ | $day$ | 0.0007 | Infusion time |
| BSA | $m^2$ | 2 | Body surface area |

For a personalization of the mathematical model, we used the longitudinal data to estimate the model parameters specified in Table 4.

Table 4: List of all personalized model parameters

| Parameter | Unit | Description |
|---|---|---|
| $k_{a1}$ | $1/day$ | Absorption rate of lenograstim |
| $k_{tr}$ | $1/day$ | Transition rate |
| $\gamma$ | – | Feedback speed of G-CSF on WBCs |
| slope | $L/\mu mol$ | PD effect of Ara-C on WBCs |
| $B$ | $10^9/L$ | Baseline of WBC count |
| $x^0_{blasts}$ | $10^9/L$ | Relative number of blasts at start of consolidation therapy |

Using a population parameter estimation (nonlinear mixed-effects modeling) approach with the software NONMEM 7.5.0, we obtained six individual model parameters for each patient. For all estimated parameters we have both patient-specific estimations and fixed effect. The fixed effects are listed in Table 5. They can be interpreted as a general prediction of the (population) parameter. The residual error was 0.199. Note that the initial values for $x_{l1} = x^0_{blasts}$ were calculated involving a heuristic function to translate the relative blast counts to absolute numbers. Furthermore, the formula $x^0_{blasts} = \hat{x} - 0.005(B\ k_{wbc})/k_{tr}$ was applied to connect estimated numbers $\hat{x}$ with another modeling assumption. See [Jost et al., 2020] for details.

Table 5: Results of the nonlinear mixed-effects modeling and numerical solution with NONMEM 7.5.0

| $B$ | $k_{tr}$ | slope | $\gamma$ | $k_a$ | $x^0_{blasts}$ |
|---|---|---|---|---|---|
| **Fixed Effect Prediction $\theta \in \mathbb{R}^6$** | | | | | |
| 4.78 | 0.202 | 6.94 | 0.707 | 4.53 | 0.00458 |
| **Interindividual Variability CV%** | | | | | |
| 37.1 | 23.6 | 32.1 | 19.8 | 81.7 | 11.4 |

We provide the covariance matrix also calculated with NONMEM 7.5.0 for the patient-specific parameter while being consistent to the order of parameters used in Table 5.

$$Cov = \begin{bmatrix} 0.137439 & 0.0268759 & 0.00550955 & -0.00340233 & -0.0792157 & 0.103258 \\ 0.0268759 & 0.055742 & -0.00304377 & -0.019963 & 0.00428422 & 0.0721956 \\ 0.00550955 & -0.00304377 & 0.102957 & 0.0259179 & -0.0780699 & 0.145333 \\ -0.00340233 & -0.019963 & 0.0259179 & 0.0392254 & -0.0321693 & 0.0107366 \\ -0.0792157 & 0.00428422 & -0.0780699 & -0.0321693 & 0.668126 & 0.101477 \\ 0.103258 & 0.0721956 & 0.145333 & 0.0107366 & 0.101477 & 1.30212 \end{bmatrix} \quad (17)$$

This covariance can be used to draw additional artificial patients based on the distribution obtained from the data available. The final parameters used for simulation are then calculated by multiplying the fixed effects $\theta$ from Table 5 with the exponential of the estimated or drawn patient-specific parameter. We denote the correspondence of a parameter to a patient or a drawn artificial patient with $i$.

$$
\begin{bmatrix} B_i \\ k_{tr_i} \\ slope_i \\ \gamma_i \\ k_{a_i} \\ x^0_{blasts_i} \end{bmatrix} = \begin{bmatrix} \theta_B \\ \theta_{k_{tr}} \\ \theta_{slope} \\ \theta_\gamma \\ \theta_{k_a} \\ \theta_{x^0_{blasts}} \end{bmatrix} \exp\left( \begin{bmatrix} \eta_{B_i} \\ \eta_{k_{tr_i}} \\ \eta_{slope_i} \\ \eta_{\gamma_i} \\ \eta_{k_{a_i}} \\ \eta_{x^0{}_{blasts_i}} \end{bmatrix} \right) \tag{18}
$$

Finally, we provide the personalized model parameters for all 65 patients we have measurements from and that were used for parameter estimation. All parameters are given with a consistent number of digits. This is not due to significance but rather due to readability and consistency with the literature values provided in Table 3.

Table 6: Patient-specific estimated model parameters

| **Patient id** | **B** | $\boldsymbol{k_{tr}}$ | $\boldsymbol{\gamma}$ | **slope** | $\mathbf{k_a}$ | $\boldsymbol{x^0_{blasts}}$ |
|---:|---|---|---|---|---|---|
| 1 | 7.9486 | 0.25944 | 0.65448 | 6.6155 | 3.2428 | 1.6867 |
| 2 | 2.8166 | 0.24445 | 0.52384 | 7.6323 | 9.0704 | 4.2797 |
| 3 | 10.592 | 0.25506 | 0.74528 | 9.6581 | 1.9137 | 4.7626 |
| 4 | 3.5361 | 0.24263 | 0.67855 | 7.5927 | 5.0817 | 2.5351 |
| 5 | 4.8955 | 0.20583 | 0.62789 | 5.4085 | 6.2395 | 1.4801 |
| 6 | 4.5859 | 0.21625 | 0.70575 | 7.5677 | 4.2518 | 1.9014 |
| 7 | 5.4688 | 0.24489 | 0.57731 | 7.2270 | 4.1366 | 1.8369 |
| 8 | 3.0416 | 0.18571 | 0.68770 | 6.3089 | 6.3185 | 1.2505 |
| 9 | 5.3121 | 0.30312 | 0.65462 | 7.4970 | 2.1573 | 5.2653 |
| 10 | 3.4184 | 0.25135 | 0.57087 | 7.4679 | 4.8991 | 1.0951 |
| 11 | 8.0523 | 0.16385 | 0.93809 | 11.590 | 1.9737 | 3.8741 |
| 12 | 2.1501 | 0.19919 | 0.67406 | 9.4983 | 4.1113 | 2.7085 |
| 13 | 5.9667 | 0.18948 | 0.65151 | 5.6163 | 4.5824 | 0.2803 |
| 14 | 3.4979 | 0.27748 | 0.54717 | 5.8753 | 8.1764 | 2.3349 |
| 15 | 4.7392 | 0.25209 | 0.56108 | 5.8816 | 5.9524 | 1.5701 |
| 16 | 5.3869 | 0.20788 | 0.60090 | 12.445 | 3.9281 | 8.8081 |
| 17 | 4.2011 | 0.23157 | 0.71396 | 6.8993 | 5.1013 | 1.0670 |
| 18 | 4.8346 | 0.28311 | 0.54539 | 6.2418 | 6.4825 | 1.2162 |
| 19 | 4.2657 | 0.19235 | 0.74215 | 5.8187 | 4.3950 | 0.4692 |
| 20 | 7.0345 | 0.25000 | 0.71213 | 5.6554 | 5.0671 | 3.7435 |
| 21 | 8.3913 | 0.25708 | 0.60111 | 4.1656 | 4.0396 | 0.4120 |
| 22 | 8.1884 | 0.30259 | 0.57469 | 6.7056 | 4.6485 | 3.3539 |
| 23 | 6.5801 | 0.23684 | 0.81094 | 8.1394 | 3.3494 | 3.5222 |
| 24 | 7.3105 | 0.25661 | 0.88452 | 9.9849 | 3.2972 | 2.2347 |
| 25 | 5.8967 | 0.13113 | 0.56008 | 7.1150 | 4.1098 | 0.3608 |
| 26 | 8.2972 | 0.19978 | 0.58588 | 6.8880 | 2.7974 | 0.4445 |
| 27 | 5.6396 | 0.15433 | 0.95078 | 9.7500 | 3.0166 | 2.9667 |

Table 6: Patient-specific estimated model parameters

| Patient id | B | $k_{tr}$ | $\gamma$ | slope | $k_a$ | $x^0_{blasts}$ |
|---|---|---|---|---|---|---|
| 28 | 2.7016 | 0.24733 | 0.55248 | 10.361 | 6.7852 | 4.2782 |
| 29 | 3.7000 | 0.21510 | 0.72975 | 9.1689 | 3.5145 | 2.3707 |
| 30 | 5.6541 | 0.23721 | 0.82594 | 10.208 | 2.2576 | 5.4032 |
| 31 | 5.2017 | 0.22406 | 0.82755 | 9.7669 | 2.8261 | 2.7652 |
| 32 | 4.0655 | 0.16873 | 0.91295 | 7.1931 | 5.4911 | 0.9803 |
| 33 | 4.7514 | 0.16954 | 0.79004 | 4.7579 | 6.5324 | 0.4055 |
| 34 | 3.4610 | 0.17505 | 0.79958 | 9.3623 | 6.3286 | 2.2810 |
| 35 | 3.4718 | 0.15294 | 1.05930 | 9.9026 | 2.4458 | 3.5586 |
| 36 | 5.2309 | 0.32344 | 0.89942 | 5.9028 | 9.3148 | 3.3537 |
| 37 | 9.5609 | 0.32892 | 0.61605 | 8.6815 | 2.2489 | 3.0188 |
| 38 | 5.0640 | 0.26599 | 0.79128 | 6.6957 | 6.0160 | 6.3722 |
| 39 | 10.139 | 0.27426 | 0.71329 | 10.003 | 3.3266 | 7.9299 |
| 40 | 5.5669 | 0.19500 | 0.64255 | 8.1875 | 3.5311 | 1.3349 |
| 41 | 4.3203 | 0.18957 | 0.79576 | 7.3622 | 4.4171 | 0.7779 |
| 42 | 4.5405 | 0.23121 | 0.60351 | 7.6093 | 4.6768 | 1.9716 |
| 43 | 4.4084 | 0.22807 | 0.67270 | 9.2018 | 4.8867 | 3.2528 |
| 44 | 5.2061 | 0.15261 | 0.79031 | 7.8732 | 3.4211 | 1.1890 |
| 45 | 4.9365 | 0.18405 | 0.64306 | 3.6496 | 11.567 | 6.2983 |
| 46 | 6.0840 | 0.17222 | 0.80826 | 4.9991 | 9.4007 | 23.698 |
| 47 | 6.2352 | 0.20943 | 0.75407 | 8.9540 | 3.2550 | 3.4045 |
| 48 | 5.1978 | 0.19175 | 0.67417 | 3.4368 | 5.3916 | 0.0021 |
| 49 | 5.4062 | 0.18606 | 0.74200 | 9.1736 | 3.5217 | 3.2048 |
| 50 | 5.6288 | 0.21284 | 0.62145 | 4.0890 | 7.5715 | 2.5029 |
| 51 | 6.6960 | 0.17720 | 0.61307 | 5.9185 | 4.2019 | 1.3424 |
| 52 | 3.9256 | 0.17039 | 0.66297 | 5.0101 | 7.2501 | 1.6524 |
| 53 | 4.8942 | 0.21438 | 0.58426 | 4.0313 | 4.9581 | 0.0394 |
| 54 | 4.2305 | 0.16222 | 1.14650 | 12.207 | 1.5185 | 0.0237 |
| 55 | 6.4670 | 0.17902 | 0.63684 | 7.4602 | 3.2723 | 1.1549 |
| 56 | 3.1446 | 0.16629 | 0.70567 | 6.7460 | 4.2348 | 0.2009 |
| 57 | 2.2922 | 0.13701 | 0.76650 | 6.6364 | 6.5720 | 0.7695 |
| 58 | 5.2768 | 0.13701 | 0.75130 | 10.684 | 2.9764 | 2.4160 |
| 59 | 3.3872 | 0.17145 | 0.78383 | 6.4929 | 5.2151 | 0.9589 |
| 60 | 5.7936 | 0.22091 | 0.76722 | 7.8194 | 3.7277 | 2.8695 |
| 61 | 3.5002 | 0.15326 | 0.69783 | 4.2491 | 7.1642 | 0.4473 |
| 62 | 3.6137 | 0.14057 | 0.78554 | 5.6245 | 5.5405 | 0.6721 |
| 63 | 8.3834 | 0.17254 | 0.78270 | 10.991 | 2.2811 | 4.1653 |
| 64 | 3.6361 | 0.17476 | 0.55405 | 6.8366 | 5.9258 | 1.6388 |
| 65 | 6.4818 | 0.14356 | 0.92028 | 6.2728 | 3.4485 | 0.9695 |

In Figure 2 **C**, **D** of the main manuscript, and Figure 2b of this supplementary material, we visualized specific trajectories using patient 8 from Table 6. To quantify the uncertainty that arises from parameter estimation we drew 100 sets of parameters from the covariance matrix 17 after scaling the matrix with the factor 0.1. To build uncertainties around the patient 8 we then multiplied the patients' parameter set from Table 6 with the exponential of the drawn parameters, so that we form the uncertainty the same way the patient parameter sets are calculated in equation 18. This way we ensure an uncertainty that corresponds to the general distribution and a reasonable range of uncertainty.

### 5 Key Performance Indicators for Leukopenia

In our numerical simulation studies we obtained stable results as manifested by the smooth and plausible distributions of values Figure 2 in the main manuscript. Thus, even if the predictions of digital twins differ from the possible outcome that a treatment would have caused in a particular patient, the stochastic consideration of a cohort seems to balance out the individual differences.

To address the issue of a comparitively small cohort of only 65 virtual patients, we **quantified uncertainty** via additional Monte-Carlo simulations that propagate the prior distribution of model parameters via the dynamical system to a posterior distribution of key performance indicators.

We created 100 "artificial patients" using the covariance matrix (17). As the results below show, the outcome for these artificial patients are qualitatively similar to those of the 65 digital twins. We do not expect that even larger virtual cohorts based on (17) that would have to be simulated with considerable computational amount would change this. When considering the size of the cohort, we would also like to point to previous work [Reimann et al., 2023] in which it could be shown that the digital cohort shows no significantly different behavior from clinical cohorts involving several hundred patients.

We evaluated three virtual cohorts which differed in the model parameter values listed in Table 4. The cohort **Digital Twins** consists of the values specified in Table 6, the cohort **Artificial Patients** uses 100 parameter vectors based on the covariance matrix and the cohort **Base Value** consists of only 1 artificial patient based on the fixed effects vector $\theta$ from Table 5. For all these cohorts we simulated 323 different schedules using 3 consecutive CCs for each simulation which sums up to 160 854 considered CCs in total.

Similar to our previous work in [Reimann et al., 2023] the goal of this study was to compare different AML treatments. Three criteria to be taken into account are leukopenia (neutropenia), leukemic blasts, and overall amount of administered drugs. For leukopenia, we mainly considered and discussed the WBC recovery time in the main article. However, our approach allows also for an easy evaluation of alternative performance indicators.

While *WBC recovery time* is defined by the number of days between the start of a CC and recovery of WBC $> 1000/\mu L$, the *leukopenia duration* is given by the number of days between the first time that WBC drop below the critical threshold and the first time they recover back $> 1000/\mu L$. This duration is by definition shorter than WBC recovery time and can be expected to be a better indicator for the time a patient is at highest risk. Our approach also allows to measure the lowest concentration of white blood cells during a consolidation Cycle. Although this does not have a clinical relevance we can consider it as a measurement of severeness of the leukopenia. Since not every simulation and in practice not every patient has a leukopenia during every CC we can also consider the percentage of simulated CCs that showed a WBC below $1000/\mu L$.

We collect the heatmaps in analogy to Figure 2A and 2B of the main manuscript in Table 7 considering all three cohorts (digital twins, artifical patients, and the base value) and the introduced performance indicators.

The results show that the PRE-G-CSF (see main manuscript) outperformed the POST-G-CSF in every category. The only performance indicator where the difference is less pronounced is the leukopenia duration. This is due to the fact that some simulations do not recover from leukopenia and therefore start the new CC still with a WBC still below $1000/\mu L$ which often yields to the *leukopenia duration* being 35 days. This occurs primarily during SIM-G-CSF schedules, as one can see in the corresponding figure in Table 7, but reduces the color range available for the other schedules. Therefore, the advantage of PRE-G-CSF is still there and visible but less pronounced in color. An earlier administration of G-CSF overlapping with Ara-C administration (SIM-G-CSF) is discouraged. There is clinical evidence for a strong increase in WBC recovery time in this case [Ortiz et al., 1993] that we could verify in our simulations. This behavior corresponds to the triangular part in the center of all heatmaps, where all simulations show worse results.

Table 7: Comparison of different performance indicators for the three cohorts Digital Twins, Artificial Patients, and the Base Value.

| Digital Twins | Artificial Patients | Base Value |
|---|---|---|

#### Mean WBC Recovery Time

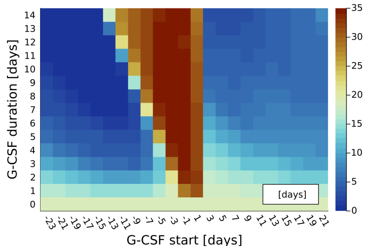

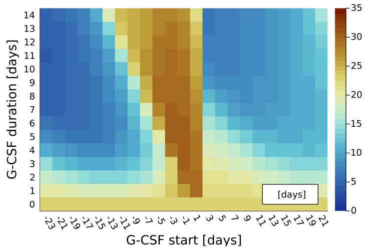

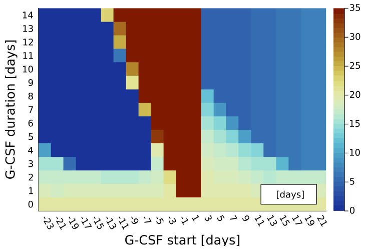


#### Mean Leukopenia Duration

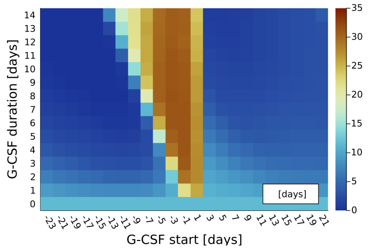

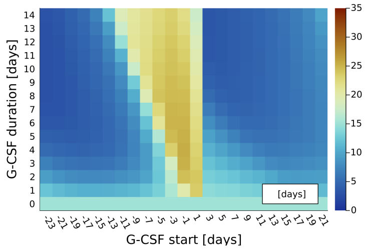

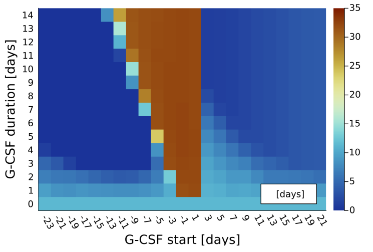


#### Mean WBC Minimum

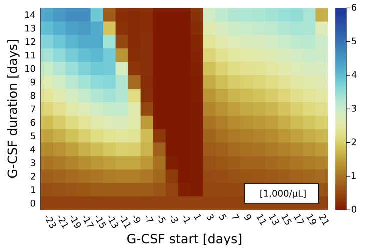

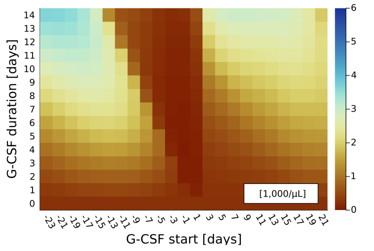

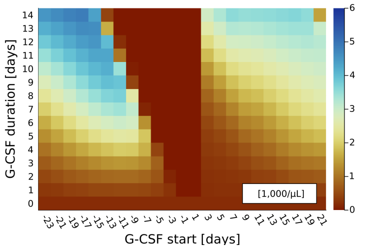


#### Leukepenia Percentage

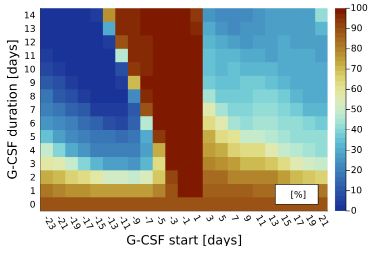

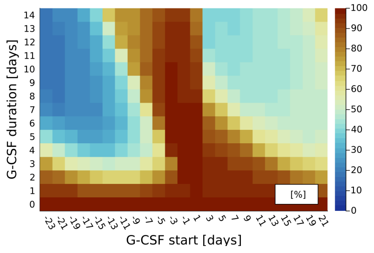

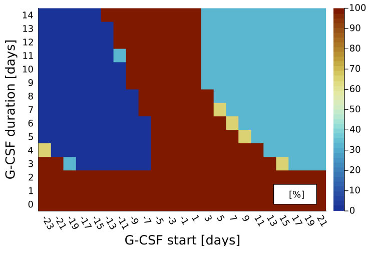

### 6 Key Performance Indicators Blasts

The second criteria we took into account when evaluating different treatments was the impact on the leukemic blasts in the bone marrow. This is very important to get a better understanding of balancing the conflicting therapy goals (avoiding severe leukopenia vs. reducing leukemic blasts).

In clinical practice it is very challenging to assess the absolute number of leukemic blasts. Bone marrow assessments are intrusive and yield uncertain relative measurements. Patients in consolidation therapy are in complete remission. In routine practice, medullary blasts in patients in complete remission are often normal immature cells. Thus, clinical decisions on how to avoid relapse have to be made without knowledge of absolute leukemic blast numbers and rely on measurable residual disease or the OS of previous clinical studies. The situation is completely different in numerical studies, because these allow an exact "observation" of the predicted values. But also in our context of training the mathematical model the sparsity and uncertainty of blast measurements was a challenge. For some of the 65 patients there were 1 or 2 measurements of BM blasts in our data. We used a heuristic measurement function to transfer the relative blast count into an absolute number [Jost et al., 2020] and estimated model parameters $x^0_{blasts}$ with this data for all the patients. For patients without available measurements the value was set to the average fixed effect or population parameter value of $x^0_{blasts}$. In summary, the initial values $x^0_{blasts}$ are very poorly approximated and uncertain. That would be a problem if we wanted to make individual decision-support or OS prediction.

For our focus on dynamics, however, this uncertainty is more or less irrelevant. By modeling the most impactful physiological processes of blast proliferation, we were able to predict absolute leukemic blast numbers for all digital twins. By considering the ratio between absolute numbers at the end and beginning of treatment, which can be easily extracted from the numerical solution, we could focus on the question of how much a treatment decreases (or increases) the absolute number of leukemic cells in comparison to other treatment choices.

This is a well-established approach in mathematical oncology. For example, the Norton-Simon hypothesis "Chemotherapy success is proportional to the growth rate of proliferating cancerous cells" and an investigation of the right hand side of the differential equation describing cancerous cell proliferation led to the insight that early, dense, high-dosage chemotherapy treatments for breast cancer are to be preferred, a triumphal success for mathematical modeling with huge clinical impact [Michor and Beal, 2015]. Norton and Simon considered Gompertz growth for the cells. Their analysis showed that timing is crucial independent of the absolute number of cancerous cells. In this line of thought we decided to analyse $\frac{x_{l1}(t)}{x_{l1}(t_0)}$, where $t_0$ marks the start of the first consolidation cycle (day 50 in this study), and to use $\frac{x_{l1}(T)}{x_{l1}(t_0)}$ where $T$ marks the endpoint of a considered consolidation cycle in time as key performance indicator kpiB.

The submodel for leukemic blasts published in [Stiehl et al., 2018] is more involved than simple Gompertz or exponential growth and considers interactions between the numbers of mature WBC and leukemic blasts. Thus, an analytical investigation is beyond the scope of this paper. Yet, the growth of leukemic cells is approximately exponential, as depicted in the supplement of [Reimann et al., 2023]. By looking at the analytical solution

$$x_{l1}(t) = x^0_{blasts}\, e^{ct}$$

of the simplified linear differential equation

$$\dot{x}_{l1}(t) = c\, x_{l1}(t), \quad x_{l1}(0) = x^0_{blasts}$$

for a constant value $c$ one observes that the ratio $\frac{x_{l1}(T)}{x^0_{blasts}} = e^{cT}$ is independent of the practically difficult to assess value $x_{l1}(0) = x^0_{blasts}$, but depends on the value $c$ which can be linked to different treatment choices. We showed that our mathematical model reproduces this behavior in section 5 of the supplementary material attached to [Reimann et al., 2023]. Thus, even if the simulated absolute leukemic blast counts are inaccurate in comparison to real patient data owing to the few available relative leukemic blast count measurements, our approach leads to a sensitivity of kpiB with respect to different treatments, but an in-sensitivity with respect to the unknown absolute leukemic blast numbers. Another advantage of this choice of kpiB is the straightforward interpretability. By design a value greater than 1 would indicate an increase of leukemic blasts, otherwise a decrease.

We are therefore interested in the question of what impact the different G-CSF schedules could potentially have on the dynamics of the absolute number of leukemic blasts. In Figure 2, we visualize mean values of the ratios of leukemic blasts in the bone marrow at the end of all CCs and the value at the start of the first CC, for all digital twins analogous to Figure 2A of the main manuscript, as well as exemplary blast simulations. As in Figure 2A and B of the main manuscript, the heatmap in Figure 2a shows the performance for 323 different G-CSF treatments. For each treatment choice the median blast ratio over 195 simulated CCs of 65 digital twins is color-encoded. One observes that blast ratios are reduced for longer G-CSF administrations (top of plot). Early administration of G-CSF (left of plot) does not perform worse than administration after chemotherapy (right of plot). Similar to Figures 2A and 2B of the main manuscript, the three classes of treatments PRE-G-CSF, SIM-G-CSF, and POST-G-CSF are clearly distinguishable by the color-encoded kpiB. While PRE-G-CSF and POST-G-CSF have a similar performance with a tendency to a reduction of leukemic blasts for increased G-CSF duration, the ratios for SIM-G-CSF are worse by a factor of approximately 2. The key insight from Figure 2a is that PRE-G-CSF does not lead to an increased risk for leukemic blast expansion/persistence as compared to POST-G-CSF.

Figure 2: Simulated impact of treatments on leukemic blast dynamics. B For the same prototypical digital twin and the three exemplary G-CSF treatments from Figure 2C and 2D, the evolution of #blasts over time is visualized.

(a) (b)

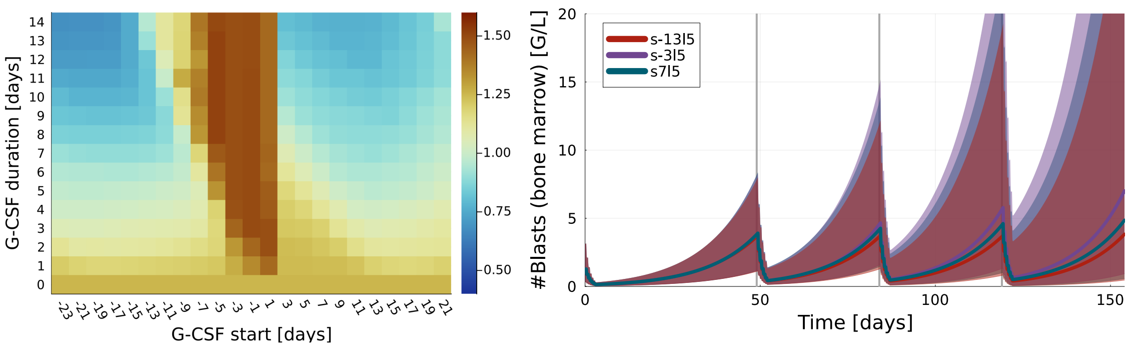


To get a general idea how the simulation of the leukemic blasts behaves we show their time evolution for three exemplary treatments of one prototypical digital twin in Figure 2b, as well as a range of uncertainty described in Section 4. The simulated concentrations used to calculate kpiB are indicated by vertical gray bars at the end of each CC and at the start of the first CC. For $s = -3$, one observes the correspondence to a blast ratio $> 1$ as an increase in leukemic blasts over time. For $s = 7$ we observe only a slight increase over time, and for $s = -13$ we can see a slight decrease of the simulated leukemic blasts which would yield a blast ratio $< 1$. Simulations with different CC durations (data not shown) indicate that while the absolute values of the ratios for each treatment may vary significantly, the relative comparison between different G-CSF treatments yields similar results (i.e., lower ratios for PRE-G-CSF and POST-G-CSF). However, on clearly sees that the range of uncertainty gets very large. This observation strengthens the importance of the result that the kpiB is insensitive to the parameters used for simulation but sensitive to the choice of treatment schedule and dosage (see supplementary material [Reimann et al., 2023]).

Summed up we concluded that the simulated impact on leukemic blasts using the PRE-G-CSF treatments also performs well. The results are superior to those for using SIM-G-CSF and no G-CSF, and are comparable to POST-G-CSF.
